# OSS-DBS v2: Simulation Platform for Biophysical Modelling of Deep Brain Stimulation

**DOI:** 10.64898/2026.10.10.26365197

**Authors:** Julius Zimmermann, Jan Philipp Payonk, Lam Vien Che, Joachim Schöberl, Till A. Dembek, Andreas Horn, Ursula van Rienen, Konstantin Butenko

## Abstract

Deep brain stimulation (DBS) is a well-established therapy for various neurological disorders. Clinical outcomes of DBS depend on electrode placement and stimulation programming, with growing evidence suggesting that targeting specific sites and networks can improve therapeutic efficacy. This targeting can be facilitated using high-accuracy, multi-modal biophysical models. In this article, we present the open-source platform OSS-DBS v2 that enables researchers and clinicians to build such biophysical models. Building upon previous work, the new version combines automated volume conductor generation, accurate and efficient electric field modelling and stimulation-driven neural simulations. OSS-DBS v2 provides an extensive library of intracranial electrodes, which can be modelled simultaneously to test complex stimulation paradigms. Its application programming interface facilitates integration of external data workflows, such as impedance measurements. Furthermore, an integration with the Lead-DBS toolbox offered a straightforward graphical user interface for stimulation volume and pathway activation modelling, further complemented by an optimization framework. OSS-DBS v2 relies entirely on open-source software, supports all major operating systems and is designed for efficient execution on consumer-grade hardware.

## 1. Introduction

Deep brain stimulation (DBS) is a clinically established method to treat symptoms in Parkinson’s disease [1], essential tremor [2], dystonia [3], epilepsy, and obsessive-compulsive disorder [4, 5]. Furthermore, it has been under investigational use for other indications such as major depression [6] and Alzheimer’s disease [7, 8]. As DBS requires surgical intervention, thorough treatment planning is paramount, including patient-specific targeting and subsequent stimulation programming to achieve the best possible outcome. In turn, effective stimulation programming requires an understanding of the mechanism of interaction between the applied electric current, i.e. which structures were stimulated, and the patient response, i.e. how and which symptoms improved. In an attempt to elucidate this mechanism and, ultimately, provide a better treatment, one can employ accurate biophysical patient-specific models of brain stimulation. Volume conductor models (VCM) based on medical imaging (MRI - magnetic resonance imaging and CT - computed tomography) are employed to solve for electric field distributions. The obtained electric fields are used to estimate the stimulation effect on the neural tissue, ranging from simple concepts of binary stimulation volumes, defined by the electric field magnitude [9] to complex pathway activation modelling (PAM) [10]. These stimulation estimates are then analysed in the context of behavioural outcomes to shed light on the relationship between stimulated tissue and symptom improvements.

A key component for patient-specific biophysical modelling of DBS is a flexible computational environment that can bring together medical imaging, different electrode models, numerical electric field solvers, and biophysical models approximating the stimulation effect on the neural level. Commercial applications such as COMSOL Multiphysics^®^ coupled with custom software have been long used for this task [11–13], but offer limited flexibility and extensibility. Optimally, the entire tool chain should be openly available to allow scaled application in the research community.

The first steps in the direction of fully open computational tools were taken by FEMfuns [14] and OSS-DBS v1 [15]. While FEMfuns focused on the volume conductor modelling, OSS-DBS provided a fully automated numerical pipeline for DBS studies [16–21]. Both packages relied on a deprecated finite element method (FEM) backend for the VCM, which became increasingly challenging to install due to a large number of dependencies. Furthermore, the FEM backend did not explicitly support complex arithmetic and curved elements for higher-order geometries, thus requiring numerically expensive approximations. Although OSS-DBS v1 already provided accurate and comprehensive biophysical modelling capabilities, it had been written in a non-modular way and was not easily extensible. In view of these gaps, we have developed the new version of the platform, further referred to as OSS-DBS v2. The primary goal was to address the aforementioned drawbacks and develop a flexible high-performance software. It is designed for both clinical researchers, configuring simulations via graphical user interface (GUI in Lead-DBS [22]), and expert users, who develop their own scripts using OSS-DBS v2 as a programming library. For clinical researchers, we demonstrate a use case of PAM-based electric current optimization for symptom-specific white matter targeting. For expert users, we describe how OSS-DBS v2 can be interfaced with measurements to permit model validation and calibration. In addition, we touch on its application for stereotactic electroencephalography (sEEG) as well as multiprotocol and multielectrode simulations. An overview of the new features introduced in OSS-DBS v2 is given in Figure 1.

**Figure 1:**
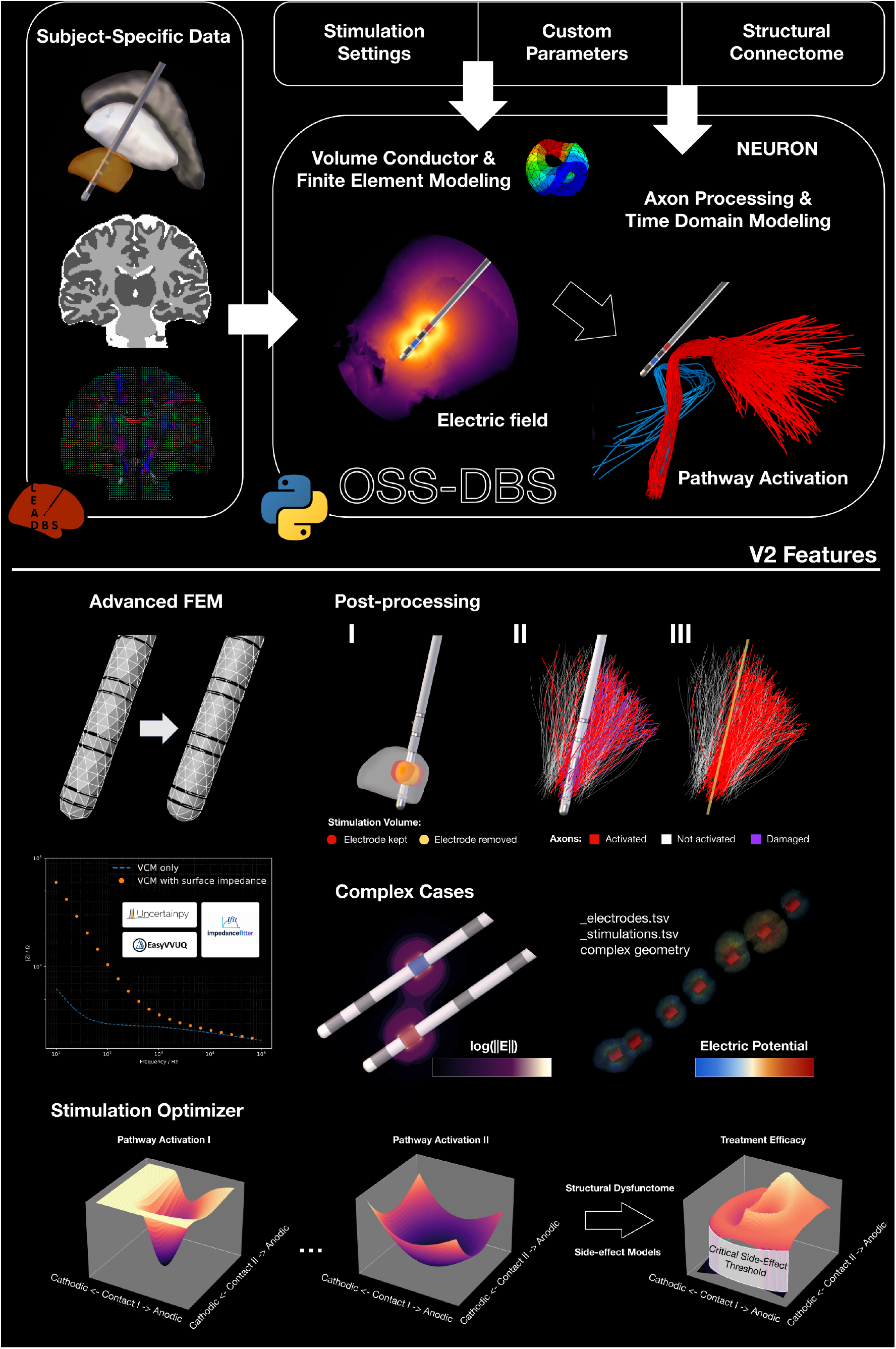
OSS-DBS v2 is a Python-based simulation library for patient-specific biophysical modelling of deep brain stimulation. Leveraging neuroimaging tools [22, 23], numerical methods [24] and neuronal models [25], it allows electric field and pathway activation modelling (PAM). The new version not only refines the originally described methodology [15], but extends it to enable new modelling paradigms. We changed the finite element method (FEM) backend to NGSolve [24], which supports curved elements, complex arithmetic, and a direct integration with CAD kernels that facilitates adding new electrode geometries. New FEM formulations unlock modelling floating potentials and impedance spectroscopy simulations. Due to the new modular framework, such simulations can now be easily interfaced with external libraries for further analysis, e.g. for uncertainty quantification. Furthermore, we introduced new postprocessing options to simulate electrode volume effect on the stimulation field and pathway activation. Beyond standard DBS problems, OSS-DBS v2 supports multi-electrode modelling and flexible coordinate-based modelling for stereotactic electroencephalogy. Finally, the newly implemented PAM-based optimizer allows effective DBS programming via targeted structural connectivity.

## 2. Methods

OSS-DBS v2 is an open-source Python library divided into multiple submodules. Individual classes and routines can be combined to write custom code in Python scripts or Jupyter notebooks. OSS-DBS v2 offers a command-line interface (CLI), which relies on an input file, and an application programming interface (API). It is supported on all major operating systems: Linux, MacOS and Windows.

### 2.1. Patient-specific modelling

#### 2.1.1. Medical imaging pre-processing

By design, OSS-DBS v2 is agnostic to imaging: users can import any pre-segmented *Nifti* data (usually with white matter, grey matter and cerebrospinal fluid labels) and define arbitrary lead coordinates in that image space. Such flexibility allows it to tackle various stimulation scenarios, including rodent DBS [18] or pre-surgical stimulation assessment. In clinical practice, tissue distribution and electrode placement are determined based on a patient’s medical imaging, i.e., MRI and CT scans. The tissue distribution is determined by segmenting the MRI data using external tools such as SPM [26], SynthSeg [27] or Atropos [28]. It is not unusual for DBS patients to have considerable brain atrophies and abnormalities that need to be accounted for by the segmentation algorithms. Import and export of *Nifti* images is handled in OSS-DBS v2 via the NiBabel library [23]. If available, information about brain tissue anisotropy can be imported as well. This information is described by diffusion tensor imaging (DTI) data stored as a 4-D array with the tensor components following the FSL ordering (xx, yx, zx, yy, zy, zz). Note that pre-processing of diffusion data is not carried out in OSS-DBS, but it can be handled by other freely available software, such as DIPY [29] and MRTrix3 [30], and converted to conductivity tensors following previously described methods [31–34].

#### 2.1.2. Geometry model

After import of the imaging and the DBS lead coordinates, the model domain and the electrodes are built using computer-aided design (CAD) modelling. By default, the modelling domain is approximated by an ellipsoid with sufficiently large dimensions to simulate remote grounding. Alternatively, users can either import a geometry from a CAD file or prepare it using the Python interface of *Netgen*[35]. The external import also permits a definition of specific boundary conditions in the VCM, e.g., electrode patches on the skull [36, 37].

OSS-DBS v2 supports most of commonly implanted DBS electrodes; for the up-to-date list, please refer to the online documentation^2^. Furthermore, a selection of sEEG- and micro-electrodes are available, as well as a tutorial on how to add new electrode models. When refactoring this module, we focused on support of multiple electrodes and simple adjustment of geometric parameters. For this, we identified key parameters of each electrode’s geometry, as specified by the manufacturer, and provided template electrode geometries with their corresponding default parameters; more details are given in the online documentation. Custom electrode geometries can be generated by providing their parameters in the input file or through the API. The outer hull of each electrode defines the encapsulation layer, which features adjustable thickness and conductivity type to enable modelling of acute and chronic implantation phases. Finally, the brain geometry, encapsulation layers and electrodes are combined into one CAD geometry. As electrodes are assumed to be perfect conductors (at the metallic contacts) or perfect insulators (remaining surface areas), their volume is excluded from the model, so their geometry is only represented by surfaces.

In OSS-DBS v2, *Netgen* [35] and its *OpenCascade* (OCC) kernel enable CAD modelling directly in the Python source code and replace *SALOME* [38] used in OSS-DBS v1. As a result, all volumes and surfaces can be labelled using a clear naming convention and directly accessed by the FEM solver *NGSolve* [24], omitting tedious communication of surface numbering and conversion of multiple mesh formats, which was necessary in the previous version.

#### 2.1.3. Mesh generation

The model geometry is discretized in *Netgen*, which features is oparametric, curved elements. DBS electrodes have rounded surfaces, and curved elements greatly reduce the geometric discretization error (see Figure 1), thereby improving the accuracy of the VCM. Furthermore, *Netgen* offers a wide range of meshing hypotheses and local mesh sizes. This allows users to leverage knowledge about physics of the problem to generate a pre-optimised mesh of adjustable quality. Examples of different meshing strategies are shown in Figure 3. For example, contact edges, where FEM singularities are to be expected due to the conductor-to-nonconductor material transitions, can be pre-refined already during the mesh generation. Alternatively, these edges can be subjected to hp-refinement, which features an automatic choice of mesh refinement *h* and polynomial order *p*. Algorithmic details about the exact implementation of hp-refinement in NG Solve can be found in [39]. Likewise, refinement can be applied to elements at brain tissue interfaces identified based on the segmented MRI and to subregions containing targeted neural tissue. These strategies can be coupled with adaptive mesh refinement (AMR) approaches, which require repeated solution of the field equation and, in our case, leverage a Zienkiewicz-Zhu error estimator. The impedance, which is computed as a volume integral over the entire solution, can serve as a stopping criterion for the AMR. For example, the mesh can be refined until the impedance does not change by more than a pre-defined tolerance.

Furthermore, material properties and boundary conditions can be mapped onto the mesh using a built-in functionality of *Netgen* and *NGSolve*. For example, the name of the brain region Brain and the encapsulation layer of the x^th^ electrode `EncapsulationLayer_X` can be used to assign specific material properties. As the mesh is encapsulated in an individual module, it could also be interfaced with FEM solvers other than NGSolve.

### 2.2. Volume conductor model

All parts described in the previous sections are ingredients for the VCM in which the electric field is numerically computed. As in the previous version, the workflow implements the Fourier FEM [40]: the stimulation signal (in time domain) is converted by a Fast Fourier Transform (FFT) to the frequency domain to solve the electroquasistaticfield equation

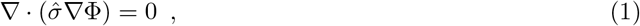

where 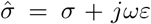 is the complex (tensor) conductivity at angular frequency *ω*. Note that the conductivity is interpolated during the FEM assembly by nearest-voxel lookup. The time domain solution is then computed using inverse Fast Fourier transform to model neural activation in response to the stimulation (see 2.3.1) [40].

Power spectra of conventional DBS signals cover a broad frequency range up to MHz. To reduce computational costs, a 250 kHz low-pass filter is applied. Still, the FFT features a linear frequency step, meaning that Eq. 1 has to be solved at thousands of frequencies. Therefore, we employ the octave band approximation as described in [41], which reduces computations to the centre frequency of the octave band and assigns the solution to all other frequencies in its band scaled by the corresponding FFT coefficients.

#### 2.2.1. Dielectric properties

Dielectric properties of brain tissue are highly uncertain [42–44]. To date, Gabriel’s model [45], which comprises four Cole-Cole dispersions, has been widely used [46]. Recently, we have suggested a correction to the model [43] that uses only three Cole-Cole dispersions and an upscaled electrical conductivity. Both models are implemented in OSS-DBS v2. In addition, the constant model is available, which uses a fixed conductivity and permittivity per material/tissue, as well as the homogeneous model to specify a scalar conductivity value for the whole computational domain. If DTI data are provided, the conductivity is represented by a tensor reflecting the anisotropic nature of brain tissue.

#### 2.2.2. Boundary conditions

OSS-DBS v2 features four types of boundary conditions (BCs): Dirichlet, Neumann, surface impedance and floating potentials. Dirichlet conditions are imposed whenever the applied voltage is known. Likewise, it is used if a known current is passed between two contacts or between a contact and the impulse generator, approximated in the model by grounding the outer surface of the computational domain. In this case, a fixed voltage drop is imposed for which the current is computed, and the solution is scaled to match the desired current. Neumann BCs are imposed on all perfectly insulating surfaces, e.g., the lead shaft. Perfectly conductive surfaces whose electric potential is not specified, e.g., passive electrode contacts, are treated as so-called floating conductors [47]. The degrees of freedom on the surface of the floating conductors are coupled into a single unknown potential value to be obtained numerically. Using this approach, a current flowing through each contact can be specified, and in OSS-DBS v2 it is used for multicontact current-controlled stimulation. OSS-DBS v2 improved here significantly over the previous version, which used a numerically challenging penalty formulation by simulating floating potentials as extremely conductive volumes.

In addition to the pure floating potential approximation, we implemented the complete electrode model, which has been shown to be in a better agreement with experimental results in the context of electrical impedance tomography [48]. The complete electrode model features a surface impedance on the floating conductor as an additional degree of freedom. We integrated *impedancefitter* [49] into OSS-DBS v2 to support arbitrary frequency-dependent surface impedance models based on equivalent circuit models.

#### 2.2.3. Numerical solver

We interfaced default solvers of *NGSolve* suitable for Eq. 1. The conjugate gradient (CG) solver is the default choice due to its speed, low memory consumption and suitability for solving the complex-valued electroquasistatic formulation (Eq. 1) in its conjugate orthogonal formulation COCG [50]. Alternatively, the Generalized minimal residual method (GMRes) [51] or a direct solver can be used. To accelerate the convergence of the iterative solvers, we precondition the linear system with a balanced domain-decomposition with constraints (BDDC) preconditioner with an algebraic multigrid (AMG) preconditioner to approximate the coarse solution as default for their low memory consumption and robust performance. For the floating conductor formulation, only the Jacobi preconditioner can be used because other preconditioners are not applicable or implemented in this setting.

### 2.3. Post-processing

At specified frequencies, the entire electric field solution, its derivatives and dielectric properties can be exported as .vtu files for visualization in ParaView (https://paraview.org/). For the estimation of stimulation volumes, the frequency domain solution at 10 kHz is used as it roughly corresponds to the median of the power spectrum of a 130 Hz 60µs rectangular pulse. All metrics to be visualized are written to comma-separated value (CSV) and Nifti files. The impedance can be computed at each frequency to probe the dielectric properties [37] and check the physical consistency of the numerical solution, e.g., by validation tests that check if the impedance is in agreement with the Kramers-Kronig relations [52, 53].

PAM requires sampling of the time domain solution along axonal trajectories. The solution at these points is saved for each frequency to an HDF5-file, and the inverse FFT is applied to retrieve the time domain solution. This reconstruction step is memory-intense: considering about 8000 frequencies and 10,000 points requires about 1.3 GB to store the computed electric potential. After it, both frequency domain and time domain solutions are available, so the memory requirement is doubled. For the given example, we would thus expect 3 GB of memory to suffice for the frequency-time conversion. Therefore, we leveraged HDF5 implementation [54] to reduce the required memory by loading the solution not at all points at the same time, but only in chunks. The data chunking, however, introduces a computational overhead and users interested in a fast processing should avoid it. It is further possible to reduce the required memory by reducing the cutoff frequency and the required number of FEM solves by checking for substantial changes in the dielectric properties (see Figure S2).

#### 2.3.1. Modelling neural response

The extracellular electric field, induced by DBS, affects various neural and non-neural tissue [55]. In turn, the neural tissue might respond to the stimulation by eliciting an action potential, further modulating the brain activity. We will refer to this phenomenon as “activation”. In general, myelinated axons are more excitable than other neural tissue [56], and the further discussion will be focused on these neural elements. Nevertheless, the concept is translatable to non-myelinated axons, dendrites and somas.

In the last years, DBS effect on various neurological and psychiatric symptoms was shown to be associated with activation of specific white matter tracts [17, 57–61] using biophysically plausible pathway activation models [10]. OSS-DBS v2 supports this approach and facilitates a setup of PAM studies via GUI and image processing implemented in Lead-DBS [22]. A typical modelling workflow is described in Figure 2.

**Figure 2:**
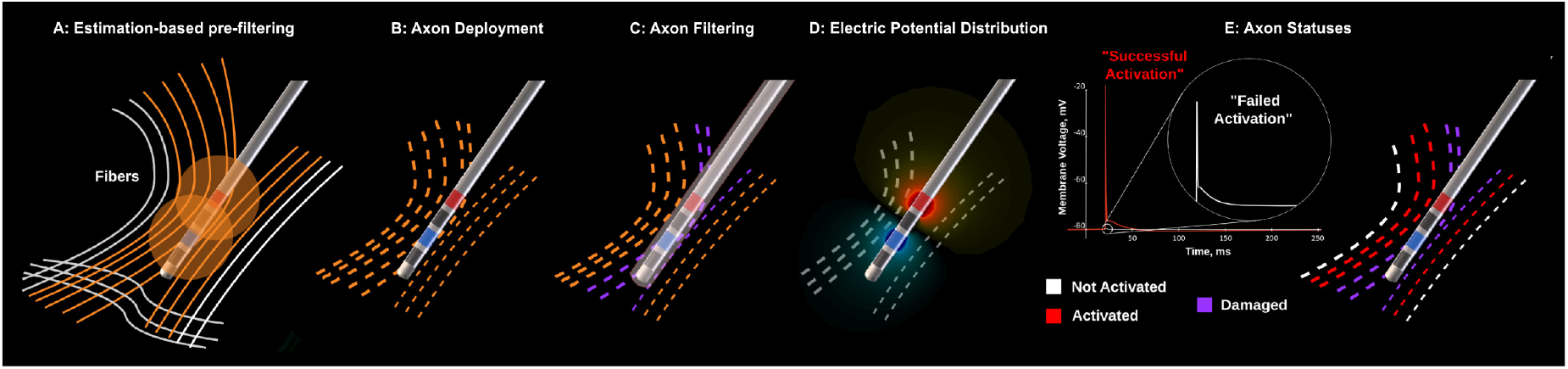
Workflow for Pathway Activation modelling. **A:**User selects a structural connectome. fibres that pass far away from active contacts are discarded using a simplified and upscaled approximation of the stimulation volume [62]. This reduces the simulation time by excluding axons where activation is highly implausible. If the modelling is conducted in native (patient) space, the remaining fibres are warped based on a precomputed nonlinear inverse transformation of patient’s MRI data to the normative space. **B:**Computational axon models are then deployed on the remaining fibres, taking into account the selected axon model and its morphology, i.e.,fibre diameter and axon length. The latter parameter refers not to the actual length of the axon, but to the model’s length along which the DBS effect is estimated. This distinction is important for long fibres, where most of the trajectory is not affected by the extracellular electric field and, hence, can be truncated to reduce computational time. One limitation of this deployment algorithm is a fixed length and diameter for fibres belonging to one pathway. **C:**OSS-DBS checks if the deployed axons are intersecting with the electrode and its encapsulation; these are marked as damaged. It also checks if the axons have unrealistic trajectories via CSF (which could be an indication of brain deficits, but also due to issues of the brain segmentation or the nonlinear transformation). In both cases, such axons are not further analyzed. **D:**OSS-DBS solves the electric field problem for the given stimulation protocol and exports the solution for the electric potential along each axon. **E:** The solution is passed to the NEURON simulator [25] as the input to the axon model. If a propagation of the action potential is detected, the axon is considered activated. Due to the linearity of the considered electric field problem, the extracellular potential can be scaled to estimate other stimulation protocols for the given volume conductor model, defined by the electrode and the surrounding tissue.

A NEURON model of an axon represents a multiple linked compartments (with discrete neural properties) at which the cable equation is solved. In OSS-DBS v2, we implemented two myelinated axon models: the “classic” McNeal’s model [63, 64] and the commonly used MRG model [65]. The latter is also available as a downsampled version, which uses an interpolated potential distribution on internodal segments, significantly reducing the memory requirements for the time domain solution. A comparison of the models is out of scope of this study, but in our experiments highly similar results were obtained across different white matter tracts for fibre diameters ranging from 2.0 to 5.0µm. It should be also noted that a selection of an appropriate fibre diameter is a non-trivial problem due to the variance reported in the neuroanatomical literature [66, 67] and a significant impact of this parameter on the activation [9, 61]. One way to address this issue is a probabilistic sampling of the diameters to obtain a probabilistic activation metric, which can inform statistical models [68].

After PAM is completed, the axons are sorted into different categories: activated, not activated and damaged (if crossing lesioned tissue, electrodes, etc). The corresponding white matter fibres, on which the axons were deployed, are then visualized in Lead-DBS (see Figure 1 and Figure 3, panels E and F). For statistical analyses, the damaged axons can be interpreted differently following strategies offered in the OSS-DBS v2 settings within Lead-DBS. Namely, such axons are either considered as activated, not activated or their status can be assessed based on the proximity to the active contacts in comparison with the activated axons.

**Figure 3:**
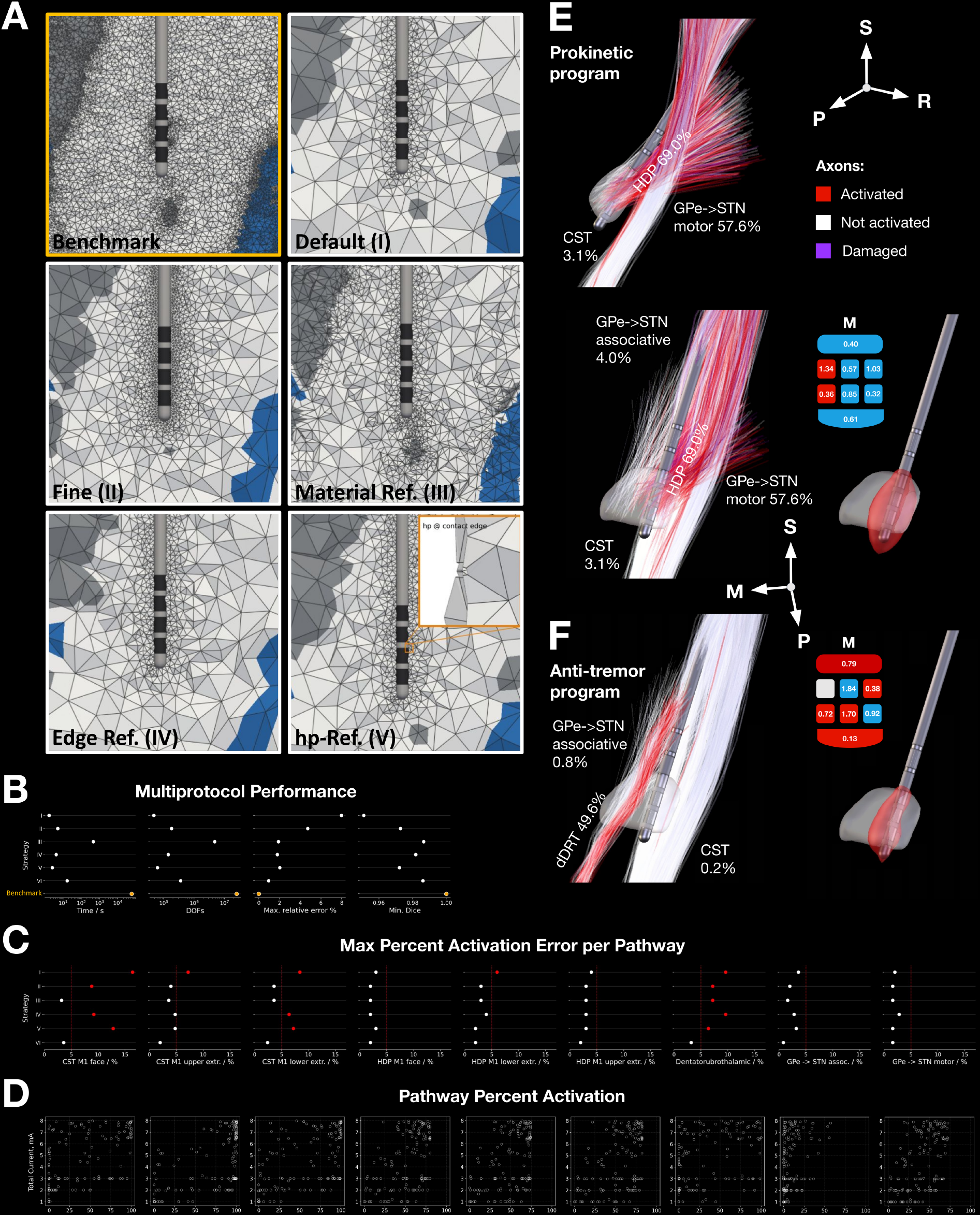
FEM error analysis of stimulation volume and pathway activation. **A:**Meshing strategies: the benchmark (see the main text), mesh generated with Netgen default preset (I), “fine” mesh preset (II), refinement at material boundaries (III) and refinement at contact edges (IV). The inset shows hp-refinement (V), which is only applied at contact edges where afield singularity is expected. Gray and white matter are shown in the corresponding colors and cerebrospinal fluid in blue. **B:**The strategies were tested on 254 stimulation protocols, containing a monopolar review and random multi contact settings. An “abstract” electrode was placed in the motor STN (see panels E and F). For each strategy, degrees of freedom (DOF) and solution time are reported as well as accuracy metrics for stimulation volumes. **C:**Maximal deviation (error) of pathway activation from the benchmark observed across the 254 stimulation protocols. The pathway trajectories were defined based on the Basal Ganglia Pathway atlas [77] The winning strategy (Fine + hp-refinement VI) was used in the subsequent optimization study. **D:**Percent activation rates of the corresponding pathways computed using the benchmark model. **E:**The “prokinetic” program aims at maximizing activation in the motor hyper direct (HDP) and pallidosubthalamic tracts (GPe-*>*STN motor), while penalizing stimulation of associative pallidosubthalamic fibres (GPe-*>*STN associative) above 5% and completely avoiding stimulation of the corticospinal tract (CST) above 5%.**F:**The same side-effect criteria are applied in the “anti-tremor” program, the targeted activation of the dentatorubrothalamic tract (dDRT) at 50%. In the figure, the actual percent activations achieved by the optimization and the corresponding currents across electrode contacts are shown (in mA; blue - cathode, red - anode). Distributions of electric field magnitudes, computed for the optimized currents and thresholded at 200 V/m, are shown on the right. It should be noted that different stimulation protocols can yield comparable pathway activations (and stimulation volumes), especially when bipolar current steering is applied.

Some steps of the workflow presented in Figure 2 can be customized. For example, users can directly provide trajectories, e.g., patient-specific tract reconstructions. It is crucial to ensure that the trajectories are defined in the same space, where the modelling is conducted, i.e., the space of the segmented MRI image. For those studies, where multiple stimulation protocols have to be evaluated, for example, for current optimization, one can leverage the linearity of the electric field problem and obtain solutions for each contact – case grounding pairs. These solutions can then be scaled and superimposed, as suggested in [69], so that only NEURON simulations have to be repeated. Note that the same approach is applicable when working with stimulation volumes based on electric-field solutions. This permits to model a new current setting in less than a second, thus facilitating optimization studies.

### 2.4. Modelling implantation effect

Implantation of a DBS electrode leads to two mechanical effects: tissue damage and tissue expansion. Currently, there is no clear consensus how these effects should be accounted for. Here, we provide two possible though not exhaustive strategies. The first strategy is to consider tissue overlapping with the electrode volume (and, optionally, its encapsulation) as damaged, see Figure 1, panels I and II.

An alternative approach is to assume that the implantation displaces neural tissue and to simulate an electrode extraction. For electric field metrics, this “pulling” the field into the electrode volume accounts for the electrode extraction. For PAM, the extraction is simulated in two steps. First, all axons overlapping with the electrode are considered activated. Second, assuming the electrode and the computational domain as a co-axial cable, we implemented the inward “pulling” by adjusting the FEM boundary conditions are adjusted, which leads to the “reduction” of the electric potential at the virtual electrode contacts:

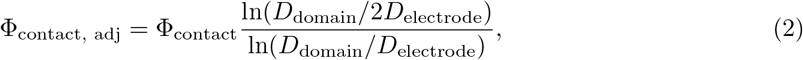

Where *D* _domain_ and *D* _electrode_ are diameters of the brain approximation and the electrode, respectively. This approach results in a more local activation in comparison with the first strategy (see Figure 1, post-processing III).

Additionally, a heuristic-based approach was implemented that computes the mean distance between the stimulation center and all activated axons. Axons overlapping with the electrode that are located closer than the mean distance are considered activated, otherwise not. The stimulation centre is defined as the amplitude-weighted average coordinate of active contacts.

### 2.5. Current optimization using pathway activation modelling

Studies utilizing PAM and its approximations identified various white matter substrates underlying improvement across different disorders and their symptoms [19, 57–61, 70–74]. In turn, these substrates can be targeted by DBS programs that take into account the patient’s clinical presentation. In this case, the stimulation current across contacts **J** is optimized to target pathways associated with alleviation of dominant symptoms while avoiding possible side-effects. Mathematically, the estimated symptom improvement *I*_symptom_ is described as

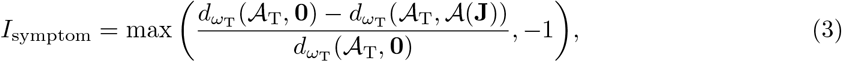

where the difference between the targeted *A*_T_ and the computed *A*(**J**) activations is assessed by a distance metric *d*_*ω* T_ (Euclidean, Manhattan, Cosine or other), defined in the pathway space with weights *ω* _T_ reflecting the importance of each pathway in the symptom modulation. This difference is quantified in relation to the null activation **0**, i.e., no stimulation case. Note that if the distance between (**J**) and *A*_T_ exceeds the distance between the latter and the null activation, *I*_symptom_ has a negative sign reflecting worsening.

The optimization algorithm of OSS-DBS v2 supports two types of side-effect thresholds. First are “soft” side-effects (SSE) that can be tolerated to some extent, but become more burdensome with increased stimulation:

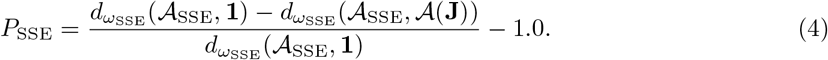

For pathways where the activation *A*(**J**) is below the corresponding threshold *A*_SSE_, their side-effect contribution (the distance) is set to 0. This distance is then compared to the full activation **1**, i.e., when all side-effect pathways are completely stimulated. The second type is critical side-effects (CSE), which occurrence should be avoided completely. The optimizer penalty *P*_CSE_ for such cases is by magnitudes larger

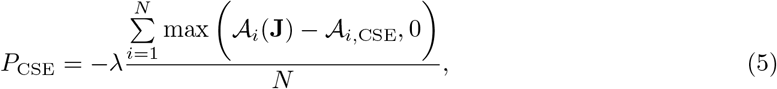

where *A*_*i*,CSE_ is the threshold for pathway *i* and *λ* is the scaling factor, e.g. 10^6^. In any case, the post-processing routine will not offer solutions with predicted critical side-effects for a clinical consideration.

Finally, the optimization problem is formulated

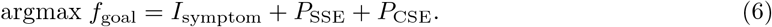

Note that multiple symptoms and side-effects can be optimized simultaneously, and their contributions can be weighted depending on patient’s primary concerns.

Optimization parameters, such as distance metrics, pathways’ weights, current limits and others are set via a GUI (PAM Optimizer) implemented in Lead-DBS. For the optimization algorithm, users can choose either Dual Annealing [75] or Particle Swarm Optimization [76], which are suitable for the complex landscape of the pathway-symptom space. During the optimization, the linearity of the electric field problem is exploited, i.e., pre-computed time domain solutions across contacts are simply scaled and superimposed according to the tested protocol and passed to the NEURON simulator to obtain percent activations *A*(**J**).

## 3. Results

### 3.1. Numerical aspects: setting the stage

We analysed the convergence behaviour of the voxel-based volume conductor modelling to ensure that the FEM mesh is fine enough to resolve the different brain regions while having as few elements as possible for computational efficiency [15]. We compared various meshing algorithms in an example case built on a segmentation of the MNI 152 asymmetric 2009b template with a standard 8-contact directional DBS lead placed on a motor STN trajectory, a four Cole-Cole dispersion model [45] and a subset of pathways from the Basal Ganglia Pathway atlas [77] (Figure 3A). The electrode position was set to the distal level at X=11.7, Y=−16.0, Z=−10.2; the proximal level at X=13.7, Y=−12.7, Z=−5.0 in the MNI 152 asymmetric 2009b space, which is in accordance with a typical motor STN implantation for PD. We used as a numerical best-case reference the solution for which the element size was limited by the voxel size and AMR was continued until the impedance changed by less than 0.1%.

The results of a preliminary investigation considering only one contact combination suggested that the stimulation volume was relatively insensitive to mesh resolution for the investigated configuration, with the default meshing approach already yielding relative differences below 5% from the benchmark stimulation volume (more details in Supplementary Material). A mixture of hp-refinement combined with refinement along material boundaries deviated by less than 1% with a moderate computational cost due to a lower number of DOFs. Unlike the stimulation volume, the PAM activations appeared to depend more on the mesh refinement strategy. Still, hp-refinement strategies performed better than any other method while keeping the number of DOFs and runtime moderate. Using the optimal mesh refinement strategy, stimulation volume estimates could be obtained in a few seconds on a conventional workstation (see Table S1). PAM analyses took less than two minutes for the considered pathways. Here, the neural modelling using NEURON took longer than the actual FEM solve and postprocessing. We chose the most promising mesh refinement approaches of the preliminary investigation to analyse the dependency of the stimulation volume and neural activation for 254 different stimulation protocols. These were composed of a monopolar review and a random sampling, including bipolar and anodic protocols, with per contact currents between −4 and 4 mA and the total current limited to 8 mA with the brain surface used as the common ground. The mesh refinement was applied for eight single contact-ground solutions that were then linearly superimposed according to the protocols.

This analysis revealed that hp-refinement applied to a “fine” mesh preset with additional refinement at material boundaries provided the best trade-off between agreement with the benchmark and computational cost among the tested strategies. As in the preliminary analysis, the stimulation volume was less sensitive to the meshing hypothesis.

Another important computational aspect is the data storage. Saving the time domain solution at many spatial points can require a large storage capacity. Given the short pulse width of the conventional 130 Hz DBS pulse, the time domain solution can be truncated without affecting the NEURON solution. We investigated the ratio between the stored time vector length and the active part of the pulse, i.e., the duration of pulse width, interpulse width, and counterpulse width. For the standard monophasic 130 Hz pulse, a duration of 30 times the pulse width, i.e., 1.2ms, was sufficient to ensure the correct estimation of the pathway activation for the 254 stimulation protocols. A duration of 20 times the pulse width was in agreement within 1% of the non-truncated reference. In turn, the required data storage of the stimulation pulses truncated to 20 times the pulse width is only about 15 % of the full stimulation signal. However, the convergence might vary depending on the stimulation and analyzed pathways, and we encourage users to explore truncation parameters using our study available in the GitHub repository as a template.

### 3.2. Target-specific programming

After evaluating meshing strategies and identifying a favourable trade-off between numerical accuracy and computational cost, we applied it to optimize DBS current protocols toward predefined target pathways while limiting activation of pathways associated with side-effects. In particular, we optimized two abstract programs: “anti-tremor”, where the aim was to achieve a moderate (50%) activation of the dentatorubrothalamic tract [72] (dDRT) and a “prokinetic” program, where we aimed to stimulate the motor hyperdirect (HDP) and pallidosubthalamic pathways. Both programs had to strictly avoid stimulation above 5% for the corticospinal tract (CST), implicated in motor side-effects [78], and a “soft” side-effect threshold of 5% was applied to fibres projecting to the associative portion of the STN due to possible neuropsychiatric side-effects [79]. Note that the target activation levels and side-effect thresholds were chosen as illustrative values to demonstrate the optimization framework rather than as clinically validated thresholds. Stimulation currents **J** were optimized in the interval of −4 to 4 mA with the total current ‖**J**‖ _1_ limited to 8 mA; multicontact and bipolar configurations were allowed. Dual annealing was selected as the optimization algorithm; differences between targeted and computed activation profiles were quantified using the Manhattan distance.

The cable equation was solved using McNeal’s axon model [63] with fibre diameters set (arbitrarily) depending on the pathway: pallidosubthalamic – 2*µ*m; hyperdirect, dentatorubrothalamic and medial lemniscus – 3*µ*; and the corticospinal fibres set to 5*µ*m. Axon lengths were chosen based on the corresponding fibre lengths, with the hyperdirect projections limited to 15 mm and passing fibres to 20 mm to decrease the computational effort. As in the previous section, we performed computations in the MNI 152 asymmetric 2009b space with the aforementioned settings. The results of the optimization and estimated stimulation volumes for the optimized currents are presented in Figure 3, panels E and F.

### 3.3. Exploring DBS models

Electric field modelling depends on various aspects such as boundary conditions, tissue dielectric properties, and electrochemical models. With OSS-DBS v2, we aimed at providing a tool that is as complete as possible regarding these modelling aspects.

The choice of boundary conditions for passive and active stimulation contacts is the first modelling step (Figure 4). Passive contacts are often considered perfect conductors and modelled as so-called floating potentials [9]. However, they lead to ringing artifacts around contact edges (see Figure 4A and [9]) due to the sharp metal-insulator interface transition. Moreover, previous research has shown that a complete electrode model needs to consider the surface impedance on the electrode to match experimental results [48]. This configuration attenuates the ringing artifacts (Figure 4B), which are completely absent when homogeneous Neumann boundary conditions are imposed (Figure 4C), representing the high-impedance limit of the complete electrode model. The floating potential and the complete electrode model allow estimating the resulting voltage on a contact and could thus be used for an experimental validation of the VCM [80]. For the bipolar configuration investigated here, the choice of the boundary condition on the passive contacts did not notably impact the stimulation volume (Figure 4A-C). It differed no more than 2% between the different boundary condition configurations. The estimated tissue impedance varied by 5% between the insulating and floating contact configuration, and by 3% between the insulating and the surface impedance configuration. This deviation could potentially be leveraged to identify the correct model assumption from impedance spectroscopy experiments.

**Figure 4:**
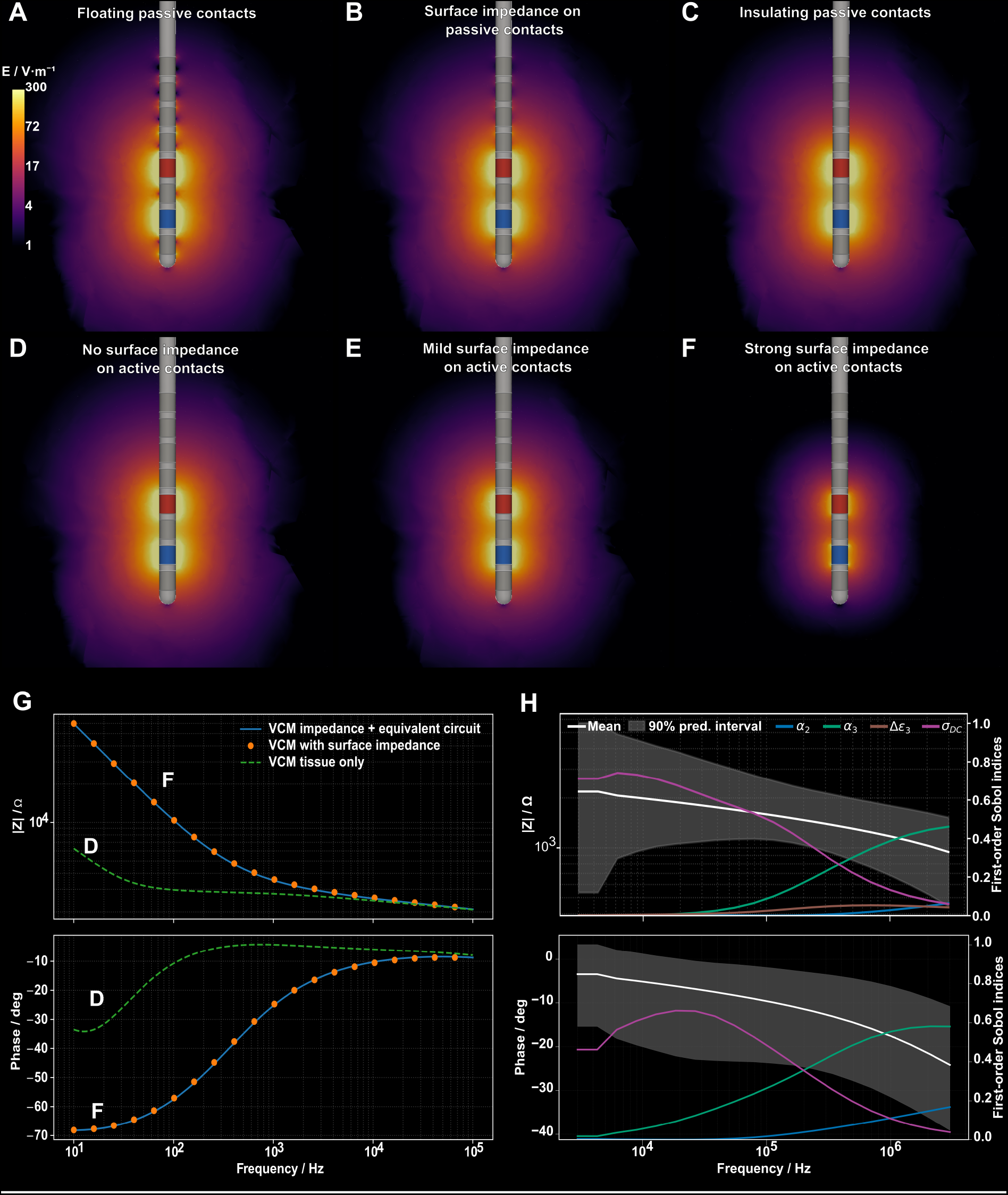
Comparison of different settings of electrochemistry boundary conditions on active (shown in red and blue) and passive contacts (shown in gray), and dielectric properties. **A** All passive contacts set to floating. Ringing is clearly visible at the contact edges. **B** An impedance of 1 kΩis imposed on the passive electrode surfaces, which already reduces the ringing. **C** The passive contacts are approximated as insulators. **D** No surface impedance is considered on the active contacts, while in **E** an impedance of 1 kΩ and in **F** an impedance of 10 kΩ was imposed. **G** Impedance modeled based on the equivalent circuit model (ECM) from [82]. The impedance of the volume conductor model (VCM) with the impedance from the ECM added to it is compared against the impedance of a VCM with a surface impedance computed from the ECM. For reference, the impedance of the tissue-only VCM is shown. **H** Uncertainty in the dielectric properties of the encapsulation tissue was propagated to quantify its influence on the real part of the impedance spectrum. Uncertainty quantification results obtained with polynomial chaos expansion using EasyVVUQ. The white solid line shows the mean impedance, and the shaded region indicates the 90 % prediction interval. The coloured curves represent the frequency-dependent first-order Sobol indices of the three-dispersion Cole-Cole model parameters of the encapsulation tissue. The broadening of the dispersion parameters *α* _2_ and *α* _3_, the static conductivity*σ* _DC_, and the dispersion strength Δ*ε* _3_ are shown. The results demonstrate that*σ* _DC_ dominates the low-frequency behaviour of the impedance spectrum, whereas *α*_3_ becomes the most influential parameter at higher frequencies. The trends are consistent with the previously reported uncertainty quantification results using polynomial chaos expansion [37].

In contrast, incorporation of the electrode-tissue interface impedance had a significant impact on the stimulation volume (Figure 4D-F). This finding is not new [81], but this aspect has so far not been considered in OSS-DBS or other open-source modelling libraries. We introduced a thinlayer Robin-like boundary condition in OSS-DBS v2 to offer this modelling option. To verify the implementation, we used the electrode-tissue interface impedance recorded and modeled by Lempka et al. [82]:the impedance of the thin-layer numerical model matched the reported superposition of interface impedance and tissue impedance (Figure 4G). Our implementation also interfaces *impedancefitter* [49], which supports arbitrary equivalent circuits. The example here serves as a verification of the surface impedance model implementation. In general, individual impedance models can be used for each stimulation contact allowing flexible modelling. This can pave the way for simulations of electrodes with electrochemically modified surfaces or significantly different surface impedances.

One challenge in DBS modelling is posed by limited knowledge of model parameters, such as the dielectric properties of brain and encapsulation tissue. Numerical uncertainty quantification (UQ) can be used to profile the DBS model with regard to model’s uncertainties. Modern UQ libraries support efficient parallel execution while reusing a precomputed computational mesh, as was demonstrated for rodent DBS models [37] using Uncertainty [83]. In the scope of this work, we demonstrated the non-intrusive integration of OSS-DBS v2 with EasyVVUQ [84], enabling the use of advanced UQ algorithms such as dimension-adaptive stochastic collocation (SC). We provide templates that are used to generate input files for OSS-DBS v2 so that the entire UQ analyses is run automatically by EasyVVUQ. Using the same parameter space of the three-dispersion Cole-Cole model for the encapsulation tissue and uncertainty ranges as in [37], the EasyVVUQ workflow reproduced the same qualitative behavior of the impedance statistics and Sobol sensitivity indices (see Figure 4).

## 4. Discussion

Over the last decade, DBS has seen multiple technological and methodological innovations. These innovations enabled increasingly complex programming strategies, for instance, due to a higher number of contacts and the possibility to activate them in arbitrary combinations with flexible electric parameters. Consequently, the programming parameter space has expanded substantially and cannot be exhaustively tested in a clinical setting. Efficient modelling of the stimulation field and its effects on neural tissue based on patient-specific or experiment-specific data enables evaluation of various settings *in silico*[19, 85], hence reducing the parameter space for *in vivo* programming. This functionality is the core of OSS-DBS, and it has been applied in both basic and clinical DBS research [16–21, 36, 37, 61, 68, 86–91].

OSS-DBS v2 provides highly performant and accurate FEM simulations for stimulation volume and pathway activation modelling. We demonstrated this through extensive convergence studies and comparison of various stimulation settings. Still, problem-specific convergence studies on a sample basis are advised because the underlying numerical models are approximate. This can, for example, be seen by the pathway-dependent sensitivity to the mesh resolution. We encourage the readers to leverage the openly shared convergence study scripts to further assess the accuracy of the different methods for problem-specific models.

Due to an efficient and memory-aware implementation of OSS-DBS v2, users can run complex biophysical models on consumer-grade hardware. Apart from the often-considered spatially homogeneous [92] and/or quasistatic models [93], the platform supports anisotropic and frequency-dependent models, integrating patient’s imaging data. Compared to other free software solutions such as StimVision [11] or ASCENT [12], which rely on COMSOL Multiphysics, OSS-DBS v2 builds upon exclusively open-source software. Consequently, this facilitates its customizability and extensibility. For example, the currently available methods to estimate neural activation can be extended by surrogate models such as driving-force predictors [94] or recently developed machine learning approaches [95].

To fully leverage the modelling potential, straightforward interoperability with external libraries for parameter optimization and uncertainty quantification was implemented [36, 37]. Previously, uncertainty quantification of DBS models required either a dedicated manual implementation [96] or modification of the simulation software [41]. Our interface removes this barrier: such methods can now be applied to OSS-DBS models without changes to its code, which makes them accessible to users without expertise in numerical methods. Future research might focus on surrogate modelling for clinical real-time applications, where OSS-DBS v2 would serve as a high-fidelity numerical reference, and on inverse modelling, i.e., calibrating the model based on various measurements.

In this work, we also addressed the question of appropriate boundary conditions, which has received attention in simulation studies [69, 97]. Here we provided boundary conditions that cover multi contact voltage- and current-controlled stimulation with appropriate numerical solvers. Once more efficient solutions are available, they can be directly integrated into the platform due to its modular and transparent implementation. For example, for the floating-conductor formulation, we currently rely on a Jacobi preconditioner, which might not be the ideal choice regarding runtime and stability. However, this choice enabled compatibility with all operating systems as we do not rely on solvers from third-party libraries besides Netgen/NGSolve. Beyond computational considerations, we also aimed to implement boundary conditions and model formulations that facilitate experimental validation of the VCM. Electrochemical impedance spectroscopy is an important tool to understand tissue properties [37] and the accuracy of electrode models [36] by comparing measured data against computed data. OSS-DBS v2 permits estimation of patient-specific impedance spectra that can be used to further calibrate model parameters of brain conductivity [98]. Likewise, the build-up of an encapsulation layer could be understood by matching computed and measured impedance spectra [37]. In general, future research should focus on integrating multimodal recordings with numerical models to enable parameter calibration. Eventually, this may close the loop between biophysical models of DBS, impedance and electrophysiological recordings. This could also enable the use of such software tools for detection and prediction of electrode failure.

Notably, OSS-DBS v2 is not limited to modelling of DBS electrodes. For example, it also offers a selection of various sEEG leads, which have comparatively longer contact spans and thus require modelling adjustments (Figure 1). Furthermore, the sEEG community often uses custom reconstruction pipelines, and therefore we implemented a support for the BIDS-standard electrode coordinate sheet. An example of sEEG field modelling is presented in Supplementary Video I. OSS-DBS v2 also allows modelling interlead stimulations, which could be informative for sEEG-based ablations and special neuromodulatory cases. A tutorial on how to set up such a simulation is presented in Supplementary Video II. Other electrodes and systems could be integrated in a similar fashion in future research. Last but not least, the development team provides user support and maintains the software including unit and integration testing, bugfix releases, and continuous updates of supported Python and third-party library versions.

### 4.1. Limitations

OSS-DBS v2 does not come without limitations, with most of them related to modelling assumptions. For example, a linear interpolation between conductivity values, which has been used in the past [99], is not yet implemented. Due to the lack of reliable measurement data to test this assumption, we have decided against it. Currently, the considered field formulation is linear, which precludes modelling of nonlinear electrochemical effects at the electrode-tissue interface [100] that have been observed in current-controlled DBS [81]. As strong electrochemical reactions represent undesired stimulation conditions, we have not focused on this aspect. Likewise, only one signal shape across all contacts is supported for the entire simulation experiment, which is in line with the current clinical practise. When using PAM, homogeneous axon parameters within one pathway are assumed, i.e., the same axon diameter and length is set for its fibres. This was an intentional choice motivated by statistical and computational considerations, and the issue was partially addressed by a probabilistic sampling [68]. The modular structure of OSS-DBS v2 permits to improve upon these assumptions when evidence for more complex models arises.

Answering the question “Which model is the correct model?” goes beyond the scope of this work. Some parameters require further experimental validation and calibration, e.g., the tissue dielectric properties [43] or the surface impedance [81]. With OSS-DBS v2, we intend to further close the gap between experiment and theory and provide a framework for validated biophysical DBS models.

## 5. Data availability

All data and code used in the study are openly available at https://github.com/SFB-ELAINE/OSS-DBSv2 and https://github.com/netstim/leaddbs/tree/develop. Online documentation can be found at https://oss-dbsv2.readthedocs.io/en/latest.

## 6. Author contributions

J.Z.: conceptualization, methodology, software, formal analysis, investigation, visualization, writing – original draft. J.P.P.: software, methodology, writing – review and editing. L.V.C.: software, investigation, visualization, writing – original draft. J.S.: software, methodology, writing – review and editing. T.A.D: methodology, formal analysis, writing – review and editing. A.H.: resources, writing – review and editing, funding acquisition. U.v.R: resources, writing – review and editing, funding acquisition, supervision, project administration. K.B.: conceptualization, methodology, formal analysis, visualization, writing – original draft, supervision.

## 7. Acknowledgements

We thank Johannes Reding, Shruthi Chakravarthy, Michael Loftus, Kenichi Maeda, Ningfei Li and Tom Reincke for their contributions to the project, and the Lead-DBS community for bug reports and feature requests.

## 8. Funding

This work was funded by the Deutsche Forschungsgemeinschaft (DFG, German Research Foundation) through SFB 1270/1,2,3 - 299150580 (J.Z, J.P.P, L.V.C, A.H., U.v.R., K.B) and CRC-1451-431549029 (A.H.). A.H. was supported by the Schilling Foundation and the National Institutes of Health (R01MH130666, 1R01NS127892-01, 2R01 MH113929 & UM1NS132358). The funders had no role in study design, data collection and analysis, decision to publish, or preparation of the manuscript.

## 9. Competing interests

T.A.D. receives research funding from Boston Scientific unrelated to this work. A.H. serves as a co-inventor on a patent granted to Charité University Medicine Berlin that covers multisymptom DBS fibre filtering and an automated DBS parameter suggestion algorithm unrelated to this work (patent #LU103178) and reports lecture fees for Boston Scientific and is a consultant for Modulight.bio. J.Z. is currently employed by Brainlab SE. The work presented in this manuscript was performed prior to his employment at Brainlab, and Brainlab had no involvement in the study, analysis, or manuscript preparation. The remaining authors report no competing interests.

