## Supplementary Information for "OSS-DBS v2: Simulation Platform for Biophysical Modelling of Deep Brain Stimulation"

*Convergence study*

First, we analysed the impact of the meshing hypothesis on the stimulation volume by applying a voltage of 1 V between two contacts and estimated the stimulation volume by thresholding the electric field magnitude at 200 V/m.We considered three contact configurations: two adjacent contacts, the two most distant contacts and the centre contact versus the grounded brain surface (case grounding). An exemplary result of the convergence study is shown in Figure S1. In general, we observed that the stimulation volume was relatively insensitive to mesh resolution for the investigated configuration, with the default meshing approach already yielding deviations below 5% from the benchmark. A mixture of edge refinement at the active contact together with refinement along material boundaries led to a deviation of about or less than 1%. This approach is a good compromise between accuracy and computational requirements. Using refinement at material boundaries together with hp-refinement slightly increased the number of DOFs while usually improving accuracy.

Additionally, we investigated the convergence of PAM models. We use the MNI 152 asymmetric 2009b template with a standard 8-contact directional DBS lead placed on a motor STN trajectory, a four Cole-Cole dispersion model [45] and a subset of pathways from the Basal Ganglia Pathway atlas [77] as described in the main manuscript. We applied a current of 1.2 mA to contact 8, while contacts 2, 3 and 4 each carried −0.4 mA, so that the injected and the returned currents sum to zero. The other contacts were set floating. The outer boundary of the computational domain was set to ground (0 V). We compared the pathway percent activation. The results suggest that the numerical model converges relatively fast (see Figure 3). For most pathways, the computed pathway activation does not change notably with the mesh. The only pathways, for which we observed differences larger than one percent point, are the "M1 upper extremities" and "cerebellothalamic" pathways. As with the stimulation volume computation, we observed that the hp-refinement strategies get closer to the reference solution than any other method while keeping number of DOFs and runtime low. Further refinement at the material boundaries usually brought the solution in better agreement with the reference solution while not leading to significant computational overhead.

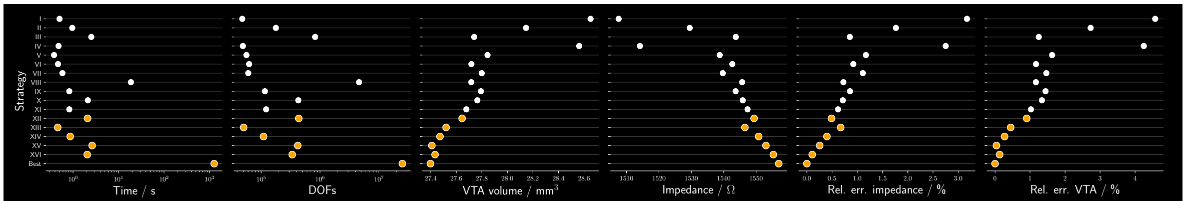


Figure S1: Comparison of mesh refinement strategies for stimulation volume estimate with case grounding. The degrees of freedom (DOF) are reported for the different strategies together with the solution time (excluding the meshing time), the stimulation volume, and the impedance. The tested strategies to approximate the best solution were: I - default meshing, II - fine meshing, III - very fine meshing, IV - edge refinement by imposing max mesh size at edges of lead perimeter divided by 20, V - edge refinement dividing the perimeter by 50, VI - edge refinement dividing the perimeter by 75, VII - edge refinement (perimeter divided by 50) and max mesh size ten times voxel size, VIII - two iterations of material refinement on very fine mesh, IX - edge refinement (perimeter divided by 50) and single material refinement, X - edge refinement (perimeter divided by 50) and double material refinement, XI - edge refinement (perimeter divided by 75) and single material refinement, XII - edge refinement (perimeter divided by 75) and double material refinement, XIII - hp-refinement, XIV - hp-refinement with single round of material refinement.

*Approximations*

Time- and frequency-domain aspects of a monophasic pulse are shown in Fig. S2. Based on this, different approximations can be applied to speed up computations. Similar considerations can be made for the neural models (Fig. S3).

*Computational benchmark*

Runtimes for various hardware and the main operating systems are shown in Table S1.

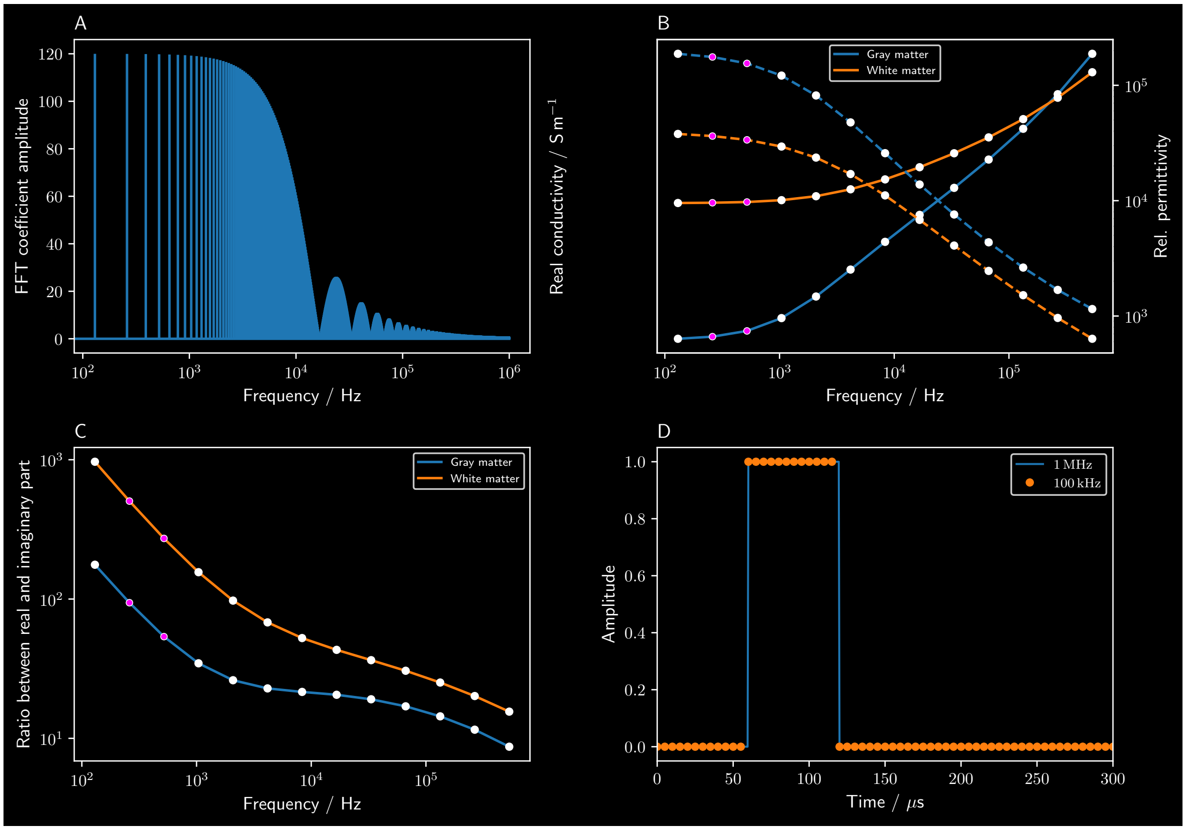


Figure S2: Different scenarios for a monophasic stimulation pulse with a pulse width of 60 µs and a frequency of 130 Hz: **A** the Fourier spectrum of the pulse includes frequencies up about 1 MHz, which means that the solution at about 7500 frequencies needs to be computed. To reduce this amount, the solution can be reused if the solution of a previous timepoint had similar dielectric properties (pink dots in **B**, here shown exemplarily for Cole-Cole-3 model [43]). In **B**, relatively few computations can be skipped, which can be improved upon by only considering the conductivity (quasistatic formulation), which is dominant at the low frequencies (**C**). The effect of the cutoff frequency is shown in panel **D**, where a reduction of the cutoff frequency to 100 kHz reduces the number of sampled points to only about 770 at the price of a reduced time resolution.

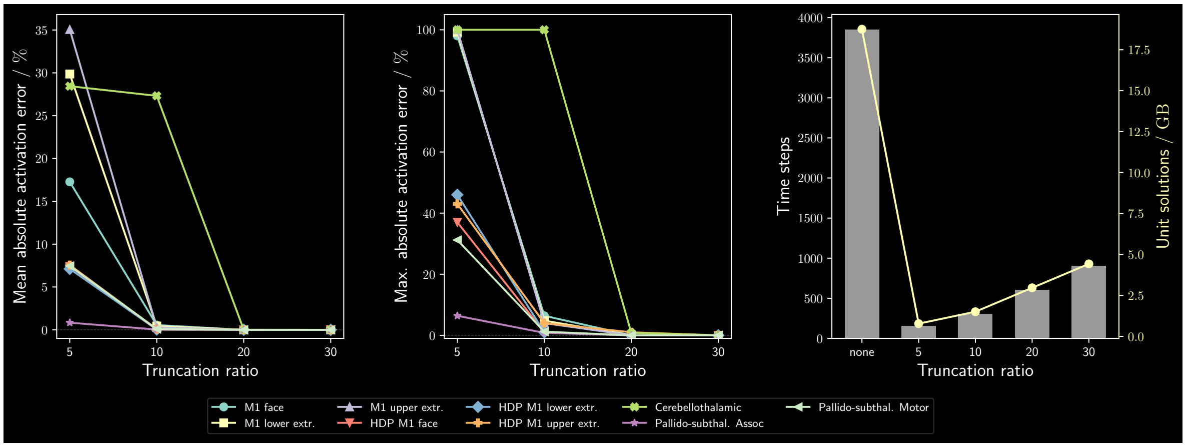


Figure S3: Effect of truncation of time domain solution and estimated neural activation. The signal was truncated at multiples of the length of the active stimulation signal, i.e., the sum of pulse width, interpulse width and counterpulse width. The truncation ratio is the proportion of the total length of the stimulation signal to its active part. The neural activation was computed for 254 stimulation protocols and the mean and max errors are shown. The right panel compares the memory consumption of the truncation levels.

Table S1: Runtime breakdown of the strategy that turned out best in the convergence study (`Fine` mesh, HP refinement with two levels and factor 0.125, one material refinement step) for different use cases. We compare PAM and stimulation volume computations with export for visualization (the lattice option) and a pure NGSolve volume computation. The fastest of three runs is reported. *Points* is the evaluation of the solution at the pathway or lattice points, *Recon.* the reconstruction of the time domain solution. *Export* refers to the time to save the computed stimulation volume to a Nifti-file. *NEURON* refers to the loading of the time domain solution and running a NEURON simulation for all considered axons. This export should be disabled if performance matters, but storing spatial information for visualization is not required.

| | | Computation steps, time in seconds | | | | | | |
|---|---|---|---|---|---|---|---|---|
| Workload | DOFs | Mesh | Solve | Points | Recon. | Export | NEURON | Total |
| *Intel Xeon E5-2687W v4 @* 3.00 GHz, *24 cores / 48 threads,* 132 GB *RAM, Ubuntu 24.04.4 LTS* | | | | | | | | |
| PAM | 357,499 | 8.3 | 19.3 | 13.7 | 10.1 | 0.8 | 41.9 | 95.7 |
| Stim. vol. (lattice) | 337,823 | 13.9 | 1.6 | 35.4 | – | 22.0 | – | 74.4 |
| Stim. vol. (NGSolve only) | 337,823 | 6.7 | 1.5 | – | – | 0.0 | – | 9.6 |
| *Intel i7-10700K @* 3.80 GHz, *8 cores / 16 threads,* 69 GB *RAM, macOS 26.6.2* | | | | | | | | |
| PAM | 357,807 | 7.9 | 25.6 | 14.3 | 6.1 | 0.4 | 49.3 | 104.8 |
| Stim. vol. (lattice) | 338,335 | 9.9 | 2.0 | 21.5 | – | 11.6 | – | 46.0 |
| Stim. vol. (NGSolve only) | 338,335 | 5.4 | 1.7 | – | – | 0.0 | – | 8.1 |
| *Intel i7-8565U @* 1.80 GHz, *4 cores / 8 threads,* 16 GB *RAM, Windows 11* | | | | | | | | |
| PAM | 355,862 | 15.9 | 55.2 | 20.9 | 13.4 | 1.0 | 143.5 | 251.1 |
| Stim. vol. (lattice) | 336,448 | 18.6 | 3.4 | 37.0 | – | 18.8 | – | 78.9 |
| Stim. vol. (NGSolve only) | 336,448 | 12.7 | 4.0 | – | – | 0.2 | – | 18.2 |
